# Parsing Genetically Influenced Risk Pathways: Genetic Loci Impact Problematic Alcohol Use Via Externalizing and Specific Risk

**DOI:** 10.1101/2021.07.20.21260861

**Authors:** Peter B. Barr, Travis T. Mallard, Sandra Sanchez-Roige, Holly E. Poore, Richard Karlsson Linnér, COGA Collaborators, Irwin D. Waldman, Abraham A. Palmer, K. Paige Harden, Danielle M. Dick

## Abstract

**Importance:** Characterizing whether genetic variants for psychiatric outcomes operate via specific versus general pathways provides more informative measures of genetic risk, and, potentially, allows us to design more targeted prevention and interventions.

**Objective:** Employ multivariate methods to tease apart variants associated with problematic alcohol use through either general or specific pathways and compare results to standard univariate genetic analysis of problematic alcohol use.

**Design:** We compared results from a univariate genome wide association study (GWAS) of problematic alcohol use to those from a previous multivariate GWAS of externalizing phenotypes. We identified genetic variants associated with problematic alcohol use through a broad liability to externalizing, and those that remain after removing shared variance with externalizing. We compared these results across SNP overlap, bioannotations, genetic correlations, and polygenic scores.

**Setting:** We included GWAS summary statistics from existing GWAS, and two US based hold out samples: The National Longitudinal Study of Adolescent to Adult Health (Add Health) and the Collaborative Study on the Genetics of Alcoholism (COGA).

**Participants:** Publicly available GWAS of externalizing behaviors and participants in Add Health (N=5,107) and COGA (N=7,483), limited to individuals of European ancestries.

**Exposure(s):** N/A

**Main Outcome(s) and Measure(s):** Outcomes included problematic alcohol use (ALCP-O), shared risk for externalizing (EXT), and problematic alcohol use-specific risk (ALCP-S) for the GWASs; a preregistered list of 99 available phenotypes for genetic correlations; and substance use, substance use disorder criteria, and alcohol misuse in the polygenic score analyses.

**Results:** The analysis differentiated SNPs operating through common versus specific risk pathways. While ALCP-O was associated with multiple phenotypes, ALCP-S was predominantly associated with alcohol use and other forms of psychopathology. Polygenic scores for ALCP-O were associated with a variety of other forms of substance use and substance use disorders, polygenic scores for ALCP-S were only associated with alcohol phenotypes. Polygenic scores for both ALCP-S and EXT show differential patterns of associations with alcohol misuse across development.

**Conclusions and Relevance:** Focusing on the differential impacts of shared and specific risk can better characterize pathways of risk for alcohol use disorders. Multivariate methods can be a useful tool for studying many psychiatric conditions. Parsing risk pathways will become increasingly relevant as genetic information is incorporated into clinical practice for psychiatric outcomes.

## Introduction

Genome-wide association studies (GWAS) are rapidly advancing our ability to detect genetic loci associated with psychiatric and substance use disorders^1–5^. However, these disorders rarely occur in isolation, and GWAS of any given outcome will detect genetic loci related to that outcome via correlated traits. This non-specificity is evident in the ubiquitous genetic correlations detected across psychiatric traits^1–5^. Alcohol use disorders (AUDs) are no exception: AUD are moderately heritable (∼50%)^6^, with most of the heritability of AUD shared with other externalizing phenotypes^7–9^, and a smaller proportion of genetic effects being specific to alcohol use outcomes^7^. New multivariate methods allow us to tease apart specific versus shared pathways by which genetic loci are associated with a given disorder^10,11^. Of particular relevance for the study of alcohol, a recent multivariate GWAS of externalizing phenotypes identified 579 loci associated with a shared genetic vulnerability to externalizing^12^. Here we apply these methods to differentiate genetic pathways for problematic alcohol use and demonstrate how moving beyond GWAS that focus on a single outcome may help us better understand *how* risk for various disorders unfolds.

Specifically, we expand upon an initial multivariate GWAS of externalizing to differentiate the genetic variants that impact problematic alcohol use through this broad externalizing liability (EXT), from variants that are specific to problematic alcohol use (ALCP-S). We compare our multivariate results to those from a previously published univariate GWAS of problematic alcohol use (the “original” GWAS results, or ALCP-O). Across these three GWAS results, we compare: 1) the genetic correlations with other relevant phenotypes; 2) the biological annotations of each genetic signal, and 3) the associations of polygenic scores (PGS) for each component with a variety of substance use phenotypes. Our analyses aim to better characterize the genetic pathways that impact problematic alcohol use and illustrate the use of multivariate genomic analyses to differentiate patterns of risk.

## Methods

### GWAS of Problematic Alcohol Use Specific Variance

In previous work^12^, we ran a common-factor, multivariate GWAS of externalizing using Genomic Structural Equation Modeling (Genomic SEM). The final model included summary statistics from seven phenotypes: (1) attention-deficit/hyperactivity disorder (ADHD), (2) problematic alcohol use (ALCP), (3) lifetime cannabis use (CANN), (4) age at first sexual intercourse (FSEX), (5) number of sexual partners (NSEX), (6) general risk tolerance (RISK), and (7) lifetime smoking initiation (SMOK). All analyses were limited to samples of European ancestry. We extended this model to simultaneously estimate the association between each single nucleotide polymorphism (SNP) and the residual variance in problematic alcohol use (ALCP-S), as, ALCP-S accounts for a large portion (∼75%) of the genetic variance (in addition to measurement error) in ALCP-O^12^.

Figure 1A displays the univariate GWAS, focusing on a single phenotype (ALCP-O). Figure 1B displays the multivariate GWAS, with the seven indicators used to estimate the model. From these two models, we derive three sets of GWAS summary statistics: the univariate GWAS results for ALCP-O (path 1), the common factor GWAS results for EXT (path 2)^12^, and the problematic alcohol use specific GWAS (path 3; ALCP-S). These three sets of results allow us to dissect genetic variance in problematic alcohol use by comparing the results of ALCP-O to both EXT and ALCP-S.

**Figure 1:**
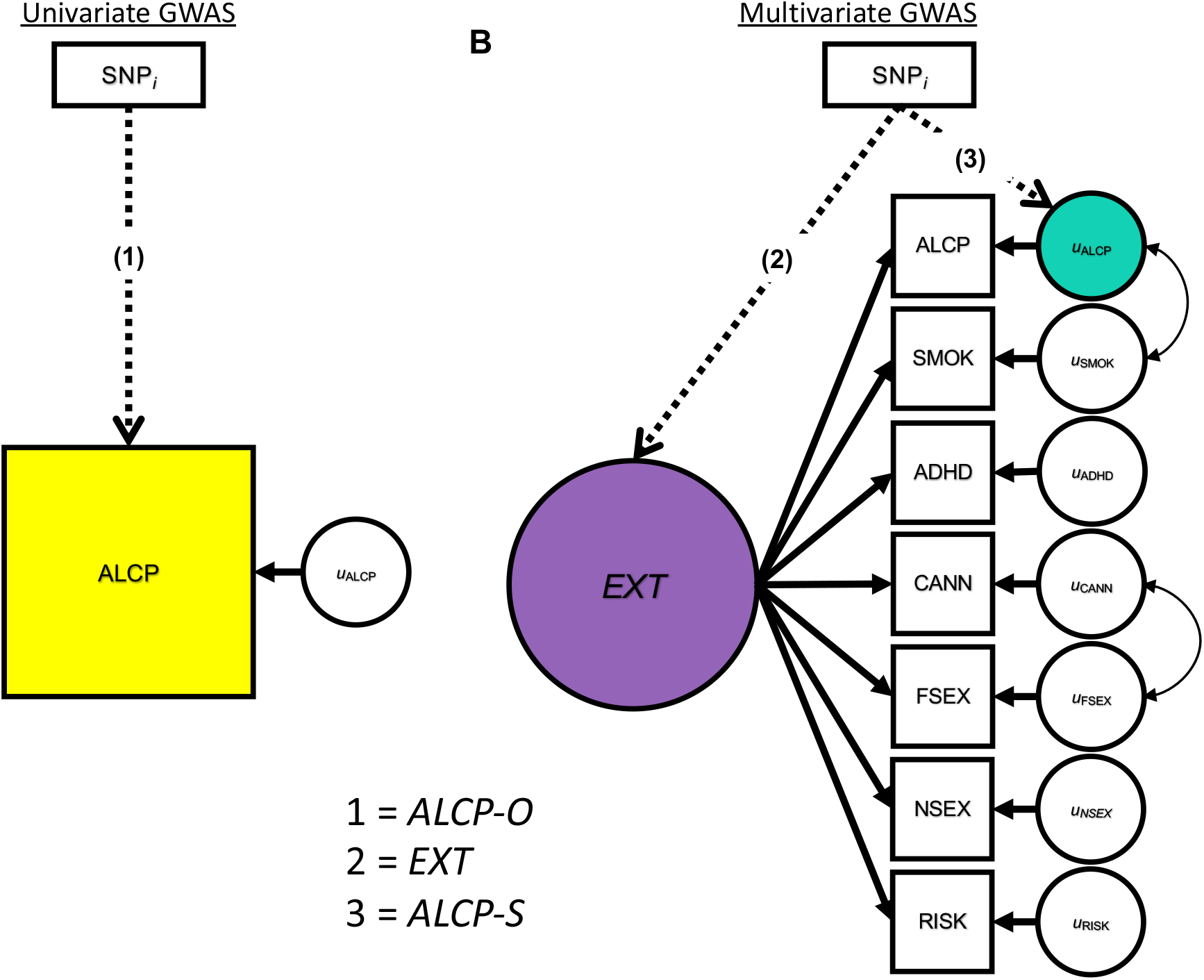
Univariate (A) and Multivariate (B) GWAS Models for Problematic Alcohol Use and Externalizing. The SNP associations with the yellow box labeled ALCP are referred to herein as “ALCP-O” (original problematic alcohol use), the SNP associations with the purple circle are referred to herein as “EXT” (shared risk towards externalizing), and the SNP associations with the teal circle are referred to herein as “ALCP-S” (problematic alcohol use-specific).

### Genetic correlations

We estimated genetic correlations between EXT, ALCP-S, ALCP-O and 99 preregistered phenotypes not included in the original model. A full list of the genetic correlations is available in Supplementary Table 2. Genetic correlations allowed us to examine how patterns of associations differed across EXT, ALCP-O, and ALCP-S. We present the change in genetic correlations from ALCP-O to ALCP-S for phenotypes that were significantly associated with ALCP-O (after correcting for an FDR^13^ of 5%).

### Bioannotation

To compare biological annotations from each of the GWAS sets (ALCP-O, ALCP-S, EXT), we performed a series of bioinformatic analyses, using a previously established pipeline^12^. First, we used FUMA^14^ v1.2.8 to identify independent SNPs (LD threshold of *r*^2^ < .1) and conduced competitive gene-set, tissue and pathway analysis using MAGMA v1.08^15^. 2) Next, we used an extension of MAGMA, Hi-C coupled MAGMA (H-MAGMA)^16^, to assign non-coding (intergenic and intronic) SNPs to genes based on their chromatin interactions. Lastly, we used S-PrediXcan v0.6.2^17^ to predict transcript abundance in 13 brain tissues, and to test whether the predicted transcripts showed divergent correlation patterns with each of the genetic factors.

### Polygenic scores

We created polygenic scores (PGS) from each set of GWAS summary statistics in European-ancestry subjects from two independent cohorts: (1) the National Longitudinal Study of Adolescent to Adult Health^18^ (Add Health; *N* = 5,107) and (2) the Collaborative Study on the Genetics of Alcoholism^19,20^ (COGA; *N* = 7,594). Within each sample we created PGS for: 1) ALCP-O; 2) EXT; and 3) ALCP-S using PRS-CS, a Bayesian approach that uses a continuous shrinkage parameter to adjust GWAS summary statistics for linkage disequilibrium (LD)^21^.

Within each of the holdout samples, we compared the effect size (incremental *R*^2^ above a covariate-only model with age, sex, and the first ten ancestral principal components) of the association of each PGS with criterion counts of AUD. Next, we compared the association of ALCP-O and ALCP-S PGS with four other substance categories, including nicotine, cannabis, opioids (COGA only), and other illicit substances (e.g., cocaine, sedatives, stimulants, methamphetamine). Substance phenotypes included “ever use” and SUD criterion counts. We corrected all analyses for multiple testing using an FDR^13^ of 5%.

Finally, we compared the association between the EXT and ALCP-S PGSs with an alcohol use index (AUI)^22^ across time in Add Health. This composite index included five alcohol phenotypes ranging from normative to problematic use, scaled to a value of 0 to 10^22^. We fit a growth model using a linear mixed model with the Add Health data structured on age. Our longitudinal analyses were limited to Add Health due to the low sample attrition across time. Detailed descriptions of the holdout samples, phenotypes, and analyses are presented in Supplementary Note 4.

## Results

### Genetic Correlations, Lead SNPs, and Bioannotations

ALCP-O had a significant genetic correlation with 64 of the 99 preregistered phenotypes. We focus on the 35 traits related to substance use, personality, and other psychiatric outcomes (full results in Supplementary Table 2). Figure 2 depicts the difference in genetic correlations (*r*_*g*_) between ALCP-O (yellow) and ALCP-S (teal), with the asterisks indicating whether a *r*_*g*_ estimate for ALCP-S remained significantly different from zero after correcting for multiple testing. Across all traits, genetic correlations from ALCP-O to ALCP-S shrank towards zero, indicating that much of the genetic variance captured in the univariate alcohol problems GWAS reflects genetic variance broadly shared with other externalizing outcomes. For substance use, ALCP-O was genetically correlated with all other forms of substance use (*r*_*g*_ = -.22 - .82). Once we remove the shared variance due to EXT, ALCP-S was only associated with drinks per week (*r*_*g*_ = .69) and age of smoking initiation (*r*_*g*_ = .14), which switched directions. As expected, ALCP-O was genetically correlated with most of the impulsivity and personality phenotypes (*r*_*g*_ = -.48 - .56). Only neuroticism (*r*_*g*_ = .33), lack of perseverance (*r*_*g*_ = .37), and positive urgency (*r*_*g*_ = .32) were correlated with ALCP-S. Finally, ALCP-O was genetically correlated with most forms of psychopathology (*r*_*g*_ = -31 - .49). ALCP-S remained associated with most of these traits, notably bipolar disorder (*r*_*g*_ = .18), major depressive disorder (*r*_*g*_ = 0.23), and schizophrenia (*r*_*g*_ = 0.16). Overall, a substantial portion of the observed genetic correlations between problematic alcohol use and other phenotypes is due to genetic variants that operate via a broad externalizing liability.

**Figure 2:**
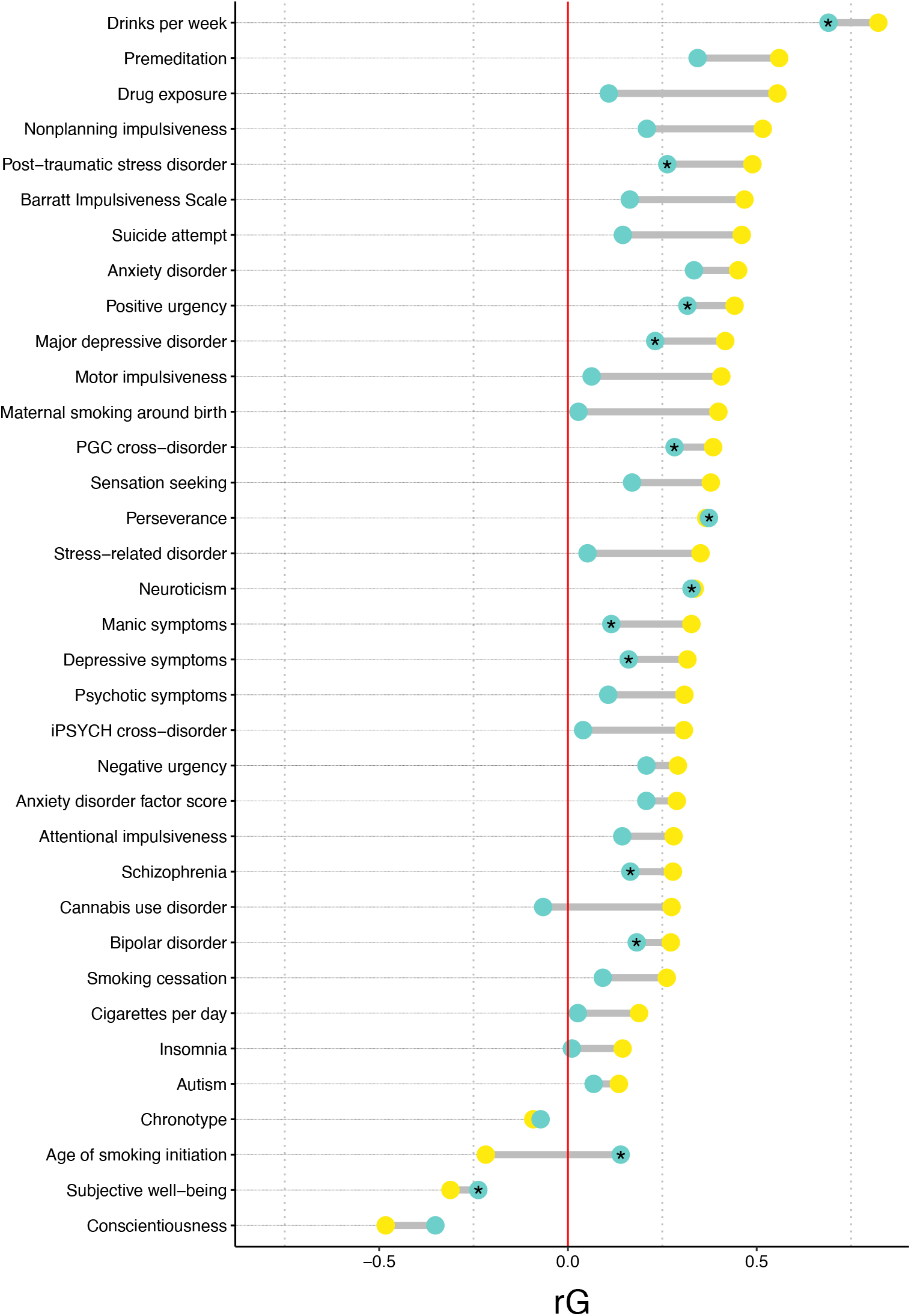
Differences in Genetic Correlations for ALCP-O and ALCP-S. Changes in genetic correlations (*r*_*g*_) from problematic alcohol use (ALCP-O, yellow dots) to problematic alcohol use-specific (ALCP-S, teal dots) across a subset of phenotypes that are significantly correlated with ALCP-O (after correcting for an FDR of 5%, **Supplementary Table 2** reports all pre-registered genetic correlations). Asterisks (*) represent genetic correlations for ALCP-S that are still significant (after correcting for an FDR of 5%).

Table 1 presents the lead SNPs from the ALCP-O GWAS across the three GWA studies. In the ALCP-O GWAS, we identified 542 genome-wide significant (*p* < 5×10^−8^) SNPs before pruning for LD. Of these 542 SNPs, 465 were genome-wide significant in EXT (∼86%) and 60 were genome-wide significant in ALCP-S (∼11%) with no overlap between these two sets. Table 1 includes the 11 independent (LD threshold of *r*^2^ < .1) lead SNPs for ALCP-O, and the comparison with the EXT and ALCP-S GWAS results. Of these, only the locus on chromosome 3 (rs10511087), in the *CADM2* region, was significant in the EXT GWAS, and the large number of SNPs before pruning were likely the result of a long-range LD region near *CADM2*. The lead SNP from another locus, located on chromosome 11, was in LD with one of the top SNPs from the EXT GWAS on *NCAM1* (rs9919558, *p* < 6.50×10^−59^). Notably, none of the SNPs on chromosome 4 were significant (or in LD) in the EXT GWAS. Instead, 8 of the 9 lead SNPs on chromosome 4 were significant in the ALCP-S GWAS. The top SNPs are in *ADH1B* and *ADH1C*, which are involved in alcohol metabolism, as well as other genes previously associated with alcohol phenotypes including *KLB*.

**Table 1:** ALCP-O Lead SNPs across ALCP-O, EXT, and ALCP-S GWASs

| SNP | CHR | BP | Nearest Gene | ALCP-O |  | EXT |  | ALCP-S |  |
| --- | --- | --- | --- | --- | --- | --- | --- | --- | --- |
|  |  |  |  | Dir | -log10(P) | Dir | -log10(P) | Dir | -log10(P) |
| rs10511087 | 3 | 85439136 | *CADM2* | + | **8.41** | + | **48.15** | + | 3.26 |
| rs6842066 | 4 | 39393801 | *RNU6-887P* | - | **9.49** | + | 0.94 | - | **10.08** |
| rs28712821 | 4 | 39413780 | *KLB* | - | **11.04** | + | 0.81 | - | **11.61** |
| rs1229984 | 4 | 100239319 | *ADH1B* | - | **46.19** | + | 1.66 | - | **48.27** |
| rs3811802 | 4 | 100244221 | *ADH1B* | - | **14.19** | + | 1.06 | - | **14.98** |
| rs3114045 | 4 | 100252560 | *ADH1C* | - | **7.88** | + | 0.67 | - | **8.30** |
| rs4699743 | 4 | 100282103 | *ADH1C* | + | **8.29** | - | 0.57 | + | **8.67** |
| rs111466094 | 4 | 100408974 | *RP11-696N14.3* | + | **8.15** | - | 2.06 | + | **9.13** |
| rs13135092 | 4 | 103198082 | *SLC39A8* | + | **14.51** | + | 1.72 | + | **13.23** |
| rs34333163 | 4 | 103283117 | *SLC39A8* | + | **7.51** | + | 1.37 | + | 6.72 |
| rs35277073 | 11 | 113350620 | *DRD2* | + | **7.49** | + | 2.47 | + | 6.40 |

Independent lead SNPs from ALCP-O GWAS pruned for an LD threshold of $r^2 < .1$ Bolded = genome wide significant ( $p < 5 \times 10^{-8}$ )

Gene-based analyses in MAGMA found 13 genes were significantly associated with ALCP-O, while only 2 genes were significant for ALCP-S. Analysis of tissue expression in MAGMA and H-MGAMA did not allow for comparisons because of the limited power in the ALCP-S results. Finally, in S-PrediXcan, only ADH1C was significantly associated with ALCP-S. While these results do not point to any new biological pathways of risk, the biological sources of genetic variance across these different pathways reaffirm that EXT is capturing a broader risk domain, while ALCP-S is largely recapitulating genes specific to the pharmacokinetics of alcohol (see Supplementary Tables 7-10 for full results).

### Polygenic Scores and Substance Use Disorders

Figure 3 illustrates associations between polygenic scores and various substance use phenotypes. Figure 3A presents the incremental *R*^2^ (Δ*R*^2^) for criterion counts in AUD. Each PGS was significantly associated with AUD criteria after correcting for multiple testing (full results in Supplementary Table 3). The ALCP-O PGS explained 0.52% of the variance 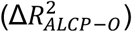 in Add Health and 1.72% of the variance in COGA. When we compare the variance explained by ALCP-O to that from adding the externalizing and the alcohol specific PGS (EXT + ALCP-S) jointly to the baseline model, we see an improvement in predictive power in both Add Health 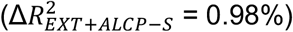 and COGA 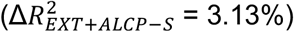.

**Figure 3:**
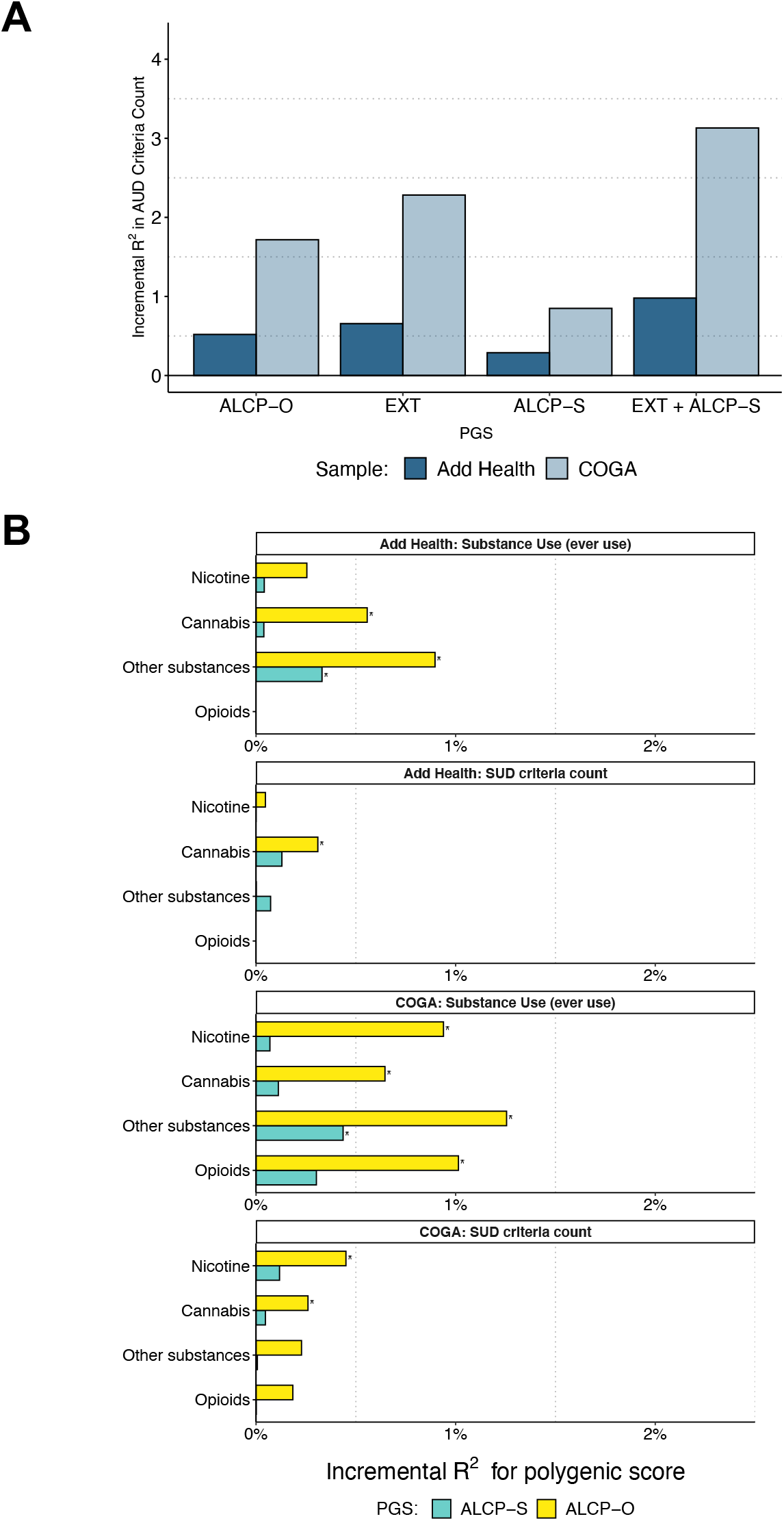
Polygenic Associations with Substance Use and Substance Use Disorders. Bar charts illustrating the incremental proportion of variance (incremental R2, or ΔR2) explained by the polygenic score in Add Health (*N* = 5,107) and COGA (*N* = 7,594). (A) Association between polygenic scores and lifetime AUD criterion counts. All polygenic scores were significantly associated with AUD criterion counts. (B) Association between polygenic scores and lifetime SUD criterion counts for cannabis, nicotine, other illicit substances, and opioids (COGA only). Asterisks (*) represent polygenic scores that are significant after correcting for an FDR of 5%.

Panel B includes other substances, in the forms of *ever use* and *SUD criterion counts*. The ALCP-O PGS is associated with cannabis use and other substance use in Add Health (OR*ALCP-O* = 1.15-1.20, Δ*R*^2^ = 0.56-0.90%) and all forms of use in COGA (OR_*ALCP-O*_ = 1.20-1.27, Δ*R*^2^ = 0.65-1.26%). Additionally, the ALCP-O PGS is associated with cannabis use disorder criteria in both COGA and Add Health (*β*_*ALCP-O*_ = 0.11-0.16, Δ*R*^2^ = 0.26-0.31%), and nicotine dependence criteria in COGA (*β*_*ALCP-O*_ = 0.20, Δ*R*^2^ = 0.45%). However, the ALCP-S PGS was only associated with ever using illicit substances (other than cannabis) and the associations are attenuated compared to that of ALCP-O (OR_*ALCP-S*_ = 1.12-1.15, Δ*R*^2^ = 0.33-0.44%). These results indicate that our multivariate analyses successfully identified variants more specifically associated with problematic alcohol use, similar to previous GWAS-by-subtraction designs^23^.

### Polygenic Scores and Longitudinal Models of Alcohol Misuse

Lastly, we fit a series of longitudinal growth models for a composite alcohol use index (AUI) in Add Health using linear mixed models (see Supplementary Note 4.5 for complete results). The best fitting model followed a quadratic change in AUI over time (with sex differences in slope), a significant association between EXT PGS and base levels of AUI (*β*_*EXT*_ = 0.12, SE_*EXT*_ = 0.01, *P*_*EXT*_ = 1.46×10^−18^), and a significant association between ALCP-S PGS and change in the linear component of age for AUI (*β*_*ALCP-S*AGE*_ = 0.07, SE_*ALCP-S*AGE*_ = 0.02, *P*_*ALCP-S*AGE*_ = 5.70×10^−5^). We found no evidence of sex-specific effects of either PGS in stratified models. Figure 4 provides a visual representation of the results from this best fitting longitudinal model across levels of each PGS (± 1.5 SD). Individuals higher on EXT PGS experience higher levels of AUI across time and sex, whereas those with higher levels of ALCP-S experience increased growth in AUI.

**Figure 4:**
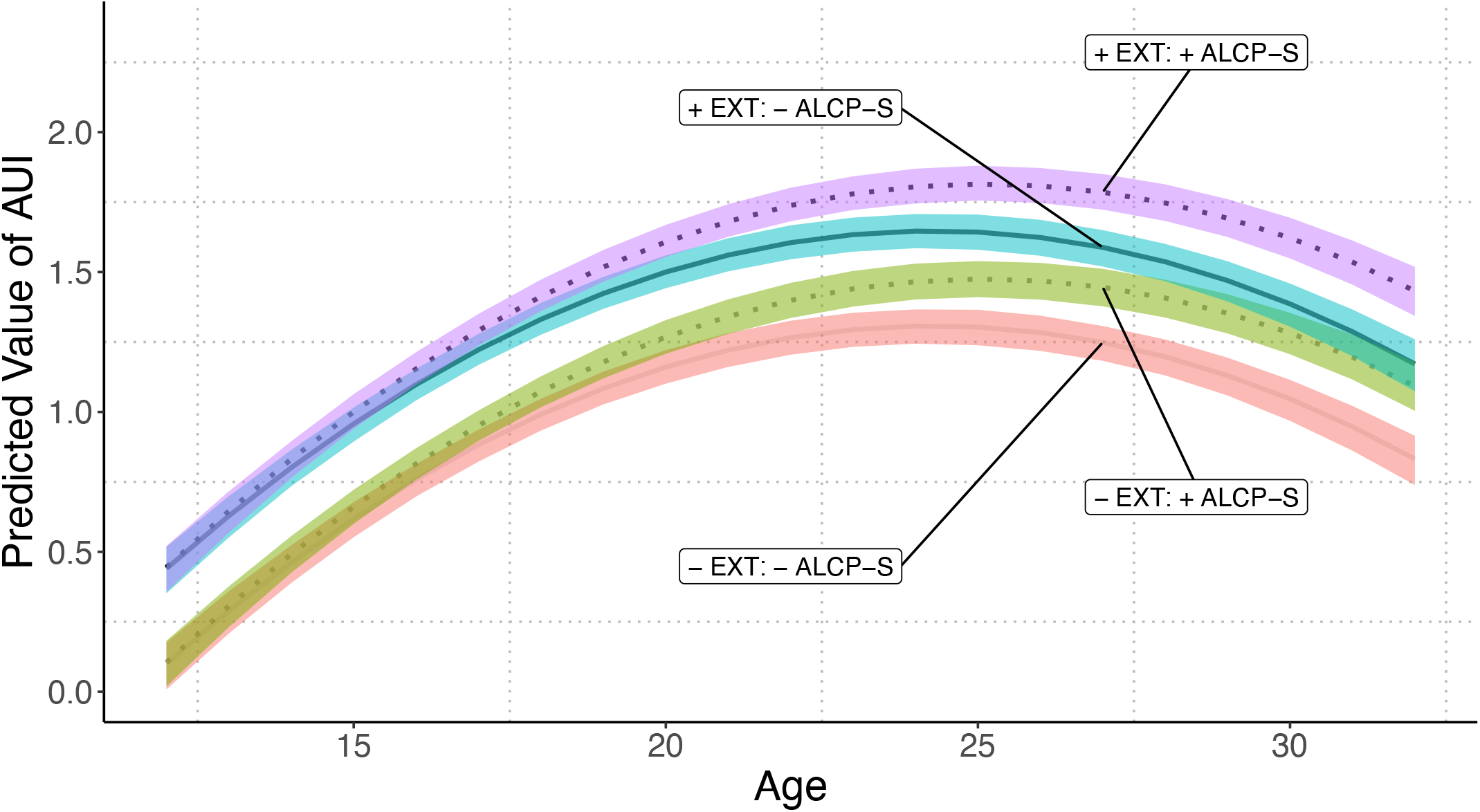
Longitudinal Models of Polygenic Associations with Alcohol Use and Misuse. Predicted values for alcohol use index (AUI) from ages 12 to 32 using linear mixed models in Add Health (N=5,107). Values for EXT and ALCP-S polygenic scores set to ±1.5 SD. The shaded areas represent 95% confidence intervals. Confidence intervals estimated using percentile method bootstrapping over 1000 bootstrap samples. All other covariates set to mean values

## Discussion

Genetic influences on SUDs can operate via both shared genetic risk with other forms of externalizing as well as via substance specific pathways. Conventional analyses focused on a single phenotype, what we would term the “classical” GWAS approach, are unable to differentiate these pathways; newer multivariate approaches begin to make this possible. In the current analysis, we demonstrated the potential of disaggregating genetic variance in problematic alcohol use into risk shared with other externalizing behaviors/disorders, and risk that is specific to problematic alcohol use. We demonstrate that multivariate genomic analyses of correlated traits can increase the specificity for characterizing *how* genetic risk unfolds.

When we compared the results from the univariate ALCP-O GWAS to those from the multivariate model (EXT and ALCP-S), we found robust evidence of distinct pathways. We compared the genetic correlations for ALCP-O and ALCP-S across 99 preregistered phenotypes, focusing here on phenotypes related to personality, substance use, and psychopathology. ALCP-O was correlated with a broad range of personality phenotypes, especially those related to impulsivity. However, after removing the variance due to the shared risk for EXT, most of these associations were no longer significant. Similarly, ALCP-O was genetically correlated with multiple forms of other substance use phenotypes, while ALCP-S remained correlated only with alcohol consumption and age of smoking initiation, and this latter correlation switched direction. The change in direction likely reflects the fact that ALCP-S seems to be capture risk for problematic alcohol use once alcohol becomes available (early adulthood), forcing the correlation with age of smoking initiation to be positive. Lastly, ALCP-O was correlated with a variety of psychiatric phenotypes. Perhaps contrary to expectation, ALCP-S continued to yield small associations with many psychiatric traits, suggesting that ALCP-S still contains signal related to other disorders. Future expansions of the multivariate model to incorporate these other psychiatric conditions would presumably be able to further tease apart the residual genetic effects on problematic alcohol use.

At the SNP level, our approach was further able to separate alcohol-specific biology from a general risk towards externalizing, given that alcohol metabolizing genes (e.g., *ADH1B* and *ADH1C*) were significant in the alcohol-specific results, but not EXT. We identified 11 lead SNPs after pruning for LD: one on chromosome 3, nine on chromosome 4, one on chromosome 11. In the ALCP-S GWAS, 8 of the 9 lead ALCP SNPs on chromosome 4 were genome-wide significant. These top SNPs were in alcohol metabolism genes (*ADH1B* and *ADH1C*), and other genes previously associated with alcohol phenotypes including *KLB*^24,25^. Additionally, *SLC39A8* has been consistently identified as a risk variant for schizophrenia^5,26^. This association with *SLC39A8 could* indicate that the ALCP-S contains variance that is not unique to alcohol (e.g., risk for internalizing or psychotic disorders). Two of these ALCP-O SNPs were in strong LD (*r*^2^ ∼ .98) with top SNPs from the EXT results. The SNP on chromosome 3 was in LD with two SNPs in the *CADM2* region, while the SNP on chromosome 11 was in LD with a SNP on *NCAM1. CADM2* has been implicated in previous GWAS of other substance use phenotypes^27,28^, risky behaviors^28–30^, and impulsivity^30^. Overall, the SNP level results broadly point to two distinct pathways of risk: one related to risk taking/impulsivity, and one specific to the body’s processing of alcohol, both of which are entwined in the univariate GWAS results for problematic alcohol use.

Finally, we evaluated polygenic scores in Add Health and COGA. Three notable findings emerge from the observed pattern of results. First, including PGS for both EXT and ALPC-S accounted for more variance in alcohol and other forms of substance use than the PGS for ALCP-O, underscoring the increase in power associated with incorporating information from genetically correlated traits^10^. Second, though the PGS from ALCP-O were associated with all forms of substance use, and SUD criteria, the PGS for ALCP-S were almost exclusively related to alcohol phenotypes, indicating that the model successfully differentiates shared and specific risk variants. Third, in longitudinal models, EXT was associated with higher mean-levels of AUI while ALCP-S was associated with increased growth in AUI. Notably, these results illustrate that externalizing genetic risk is associated with differences in AUI early in development. In contrast, during emerging adulthood, when alcohol use becomes legal and more readily accessible, there is further differentiation by alcohol-specific genetic risk. Therefore, alcohol-specific risk does not lead to alcohol problems without exposure to drinking, while broader externalizing risk captures propensity to drinking exposure across the life course. This longitudinal model reiterates the developmentally contextual nature of risk and recapitulates findings from prior studies using inferred measures of genetic risk from family-based designs^31,32^. Overall, the PGS results support the notion of a shared externalizing risk pathway and an alcohol-specific risk pathway.

Our analyses included several important limitations. First, they were limited to GWAS of European ancestries only. Unfortunately, Genomic SEM requires large sample sizes to obtain stable estimates of SNP-based heritability and genetic correlations. As larger sample sizes become available in more diverse ancestries, we will extend these models to those populations. Second, while we considered externalizing phenotypes, we did not include internalizing or psychotic disorders, which also show genetic overlap with AUD^33,34^. Finally, our estimates of SNPs associated with ALCP-S were limited by the relatively small discovery sample size for ALCP-O (N∼150K). Future iterations with more powerful GWAS of problematic alcohol^3^ use may reveal additional variants associated with ALCP-S.

GWAS of psychiatric disorders contain a mixture of different signals. Moving beyond univariate GWAS to multivariate designs offers the potential to tease apart these signals. We decomposed the genetic variation of problematic alcohol use into that which is shared with other externalizing phenotypes and that which is specific to problematic alcohol use. Comparison of results at multiple levels showed that variance specific to problematic alcohol use was related to alcohol phenotypes while that which was shared was more strongly related to other substance use and impulsivity. Differentiating these pathways of risk will become more important as genetic data becomes incorporated into clinical practice and we move towards an era of precision medicine.

## Data Availability

The full set of externalizing GWAS summary statistics can be made available to qualified investigators who enter into an agreement with 23andMe that protects participant confidentiality. Once the request has been approved by 23andMe, a representative of the Externalizing Consortium can share the full set of summary statistics. All code necessary to replicate this study is available upon request.

https://www.externalizing.org/

## Acknowledgments

**The Externalizing Consortium: Principal Investigators:** Danielle M. Dick, Philipp Koellinger, K. Paige Harden, Abraham A. Palmer.

## Lead Analysts

Richard Karlsson Linnér, Travis T. Mallard, Peter B. Barr, Sandra Sanchez-Roige.

## Significant Contributors

Irwin D. Waldman. The Externalizing Consortium has been supported by the National Institute on Alcohol Abuse and Alcoholism (R01AA015416 - administrative supplement), and the National Institute on Drug Abuse (R01DA050721). Additional funding for investigator effort has been provided by K02AA018755, U10AA008401, P50AA022537, as well as a European Research Council Consolidator Grant (647648 EdGe to Koellinger). The content is solely the responsibility of the authors and does not necessarily represent the official views of the above funding bodies. The Externalizing Consortium would like to thank the following groups for making the research possible: Add Health, Vanderbilt University Medical Center’s BioVU, Collaborative Study on the Genetics of Alcoholism (COGA), the Psychiatric Genomics Consortium’s Substance Use Disorders working group, UK10K Consortium, UK Biobank, and Philadelphia Neurodevelopmental Cohort. We would also like to thank the research participants and employees of 23andMe, Inc. for making this work possible. The full set of externalizing GWAS summary statistics can be made available to qualified investigators who enter into an agreement with 23andMe that protects participant confidentiality. Once the request has been approved by 23andMe, a representative of the Externalizing Consortium can share the full set of summary statistics. All code necessary to replicate this study is available upon request.

