## Supplemental Information for "Parsing Genetically Influenced Risk Pathways: Genetic Loci Impact Problematic Alcohol Use Via Externalizing and Specific Risk"

**Table of contents**

*Supplementary Information* ... 1
1 *Study introduction* ... 3
2 *GWAS* ... 4
2.1 **Methods** ... 4
2.1.1 Phenotype definition ... 4
2.1.2 Univariate genome-wide association analyses ... 4
2.1.3 Multivariate genome-wide association analyses ... 4
2.2 **Results** ... 5
2.3 **Genetic Correlations** ... 5
3 *Bioannotation* ... 7
3.1 **Methods** ... 7
3.1.1 Functional mapping and annotation with FUMA ... 7
3.1.2 Gene-based, gene-set, and gene-property analyses with MAGMA ... 7
3.1.3 Gene-based association using transcriptomic data with S-PrediXcan ... 8
3.2 **Results** ... 8
3.2.1 Results from the functional mapping and annotation with FUMA ... 8
3.2.2 Results from the analyses with MAGMA, H-MAGMA, and S-PrediXcan ... 8
4 *Polygenic Scores* ... 12
4.1 **Samples** ... 12
4.1.1 The National Longitudinal Study of Adolescent to Adult Health (Add Health) ... 12
4.1.2 The Collaborative Study on the Genetics of Alcoholism (COGA) ... 12
4.2 **Polygenic Score Creation** ... 12
4.2.1 Adjustment of GWAS effect sizes for linkage disequilibrium (LD) ... 12
4.2.2 Polygenic scores ... 13
4.2.3 Substance Use/Substance Use Disorder (SUD) Analyses ... 13
4.2.4 Longitudinal Growth Models ... 14
4.3 **Phenotype definitions** ... 15
4.3.1 Substance use ... 15
4.3.2 Substance use disorders ... 15
4.3.3 Alcohol Use Index (AUI) ... 16
4.4 **Results of SUD analyses in Add Health and COGA** ... 17
4.4.1 Ever Substance Use ... 17
4.4.1 Substance Use Disorder Criterion Count ... 18
4.5 **Results of Longitudinal Growth Models in Add Health** ... 18
4.5.1 Initial model building ... 18
4.5.2 Longitudinal Models of Polygenic Risk and Alcohol Misuse ... 19
5 *References* ... 21

### 1 Study introduction

Psychiatric disorders rarely occur in isolation, as evidenced by substantial cooccurrence among many psychiatric conditions. In addition to co-occurrence at the phenotypic level, there is also evidence of widespread genetic overlap across these disorders. Virtually all psychiatric disorders, including mood disorders[1], anxiety disorders[2,3], substance use disorders[4–6], and other forms of severe mental illness[7,8] are correlated to some degree at the genetic level.

While informative, these genetic correlations do not reveal the pathways through which risk for certain disorders unfolds. Individual genome wide association study (GWAS) signals contain a mixture of variants that influence the outcome through different pathways of risk. Here we highlight how new multivariate genomic methods can help us tease apart these pathways of risk, using the overlap between problematic alcohol use and other externalizing problems as an example.

Twin and family studies estimate the heritability of alcohol use disorder (AUD) to be approximately 50%[9]. A large portion (~75%) of the heritability in AUD is shared with other psychiatric and behavioral phenotypes related to externalizing. Externalizing refers to a general liability towards behavioral disinhibition and dysregulation. Externalizing includes disorders such as attention-deficit/hyperactivity disorder (ADHD) and conduct disorder in youth[10,11], substance use and antisocial behavior in adults[12,13], and normal range personality traits, such as risk tolerance, and sensation seeking[14,15]. Multiple studies have examined the phenotypic[12,15–17] and etiological[13,14,18–24] overlap between externalizing traits.

In the current analysis, we conduct a multivariate GWAS to capture genetic variance specific to problem alcohol use and demonstrate the utility in identifying genetic variance unique to a trait of interest. We do so by building upon a prior multivariate GWAS of externalizing phenotypes[24], composed of (1) attention-deficit/hyperactivity disorder (ADHD), (2) problem alcohol use (ALCP), (3) lifetime cannabis use (CANN), (4) age at first sexual intercourse (FSEX), (5) number of sexual partners (NSEX), (6) general risk tolerance (RISK), and (7) lifetime smoking initiation (SMOK). GWAS of residual phenotypes capture only a portion of the total genetic architecture of a target trait, resulting in a more specific signal. This allows for direct tests of pleiotropy, identification of specific associated variants and biological processes leading to disease, and comparison of the influence of general and specific risk pathways. The current study was performed according to a preregistered analysis plan, the first version of which was time-stamped on November 8, 2018 (https://doi.org/10.17605/OSF.IO/XKV36).

### 2 GWAS

In this section, we briefly describe the methods used to conduct the genome-wide association analyses reported in the present paper – with an emphasis on the ALCP-O and ALCP-S phenotypes. A thorough description of these genome-wide association analyses can also be found in our previously published study[24].

#### 2.1 Methods

##### *2.1.1 Phenotype definition*

In the present study, we sought to examine the general and specific genetic influences on 'problematic alcohol use'. We operationalized this phenotype as a combination of two GWASs: (1) a dimensional measure of problems related to alcohol consumption, and (2) a categorical measure of alcohol dependence. The former, a GWAS of the Problems subscale of the Alcohol Use Disorder Identification Test (AUDIT-P)[25], was analyzed in a subset of the UK Biobank (UKB) ($N$ = 130,999)[26]. The latter, a GWAS of alcohol dependence, was conducted by the Psychiatric Genomics Consortium (PGC) ($N$ = 33,685 after excluding our follow-up study cohorts)[4]. Given the large genetic correlation between these two phenotypes ($r_g$ = 0.79), results were meta-analyzed to generate a single GWAS of problematic alcohol use.

##### *2.1.2 Univariate genome-wide association analyses*

We used BOLT-LMM[27,28] v2.3.2 to conduct univariate genome-wide association analyses of AUDIT-P in UKB. As BOLT-LMM accounts for genetic relatedness among participants via a genetic variance component (pseudo $h^2$), its linear mixed model approach offers increased statistical power via the inclusion of related participants. Here, we estimated the genetic variance component using a set of 483,680 typed autosomal single-nucleotide polymorphisms (SNPs) that (i) passed the quality control procedures employed by the original UKB investigators[29], (ii) had a minor allele frequency (MAF) greater than 0.005, (iii) had a Hardy-Weinberg-Equilibrium (HWE) $P$ value greater than $1\times10^{-16}$, and light LD-pruning (window size = 50 kb; variant step-size = 5; $r^2 \geq 0.9$). Participants were excluded from analyses if they (i) did not pass the sample-level quality control thresholds employed by the original UKB investigators[29]; (ii) were part of the UKB holdout sample (or were related to those participants as indicated by a pairwise KING coefficient ≥ 0.0442); (iii) did not self-report their ethnicity as "White", "White British", "White Irish", or "Any other white background"; (iv) had discordant self-reported and genetic sex; (v) had putative sex chromosome aneuploidy; or (vi) had missing data with respect to the outcome or covariates.

We analyzed the third release of the UKB imputed genotype data. As previously reported, we included the following variables as covariates in the association analyses: sex, birth year, sex-by-birth year interactions, genotyping batch, and the first 40 genetic principal components of ancestry (see Linnér et al. 2021[24] for further detail). GWAS summary statistics were subsequently subjected to quality control procedures as previously described[24]. Finally, we used METAL[30] to conduct a sample-size-weighted meta-analysis of our AUDIT-P GWAS and the PGC's alcohol dependence GWAS.

##### *2.1.3 Multivariate genome-wide association analyses*

As we have previously described, we used Genomic Structural Equation Modeling (Genomic SEM)[31] to analyze the joint genetic architecture of externalizing phenotypes (see Linnér et al. 2021[24] for additional detail). The final confirmatory factor model included summary statistics from seven externalizing phenotypes: ADHD, ALCP, CANN, FSEX, NSEX, RISK, and SMOK. After identifying the best-fitting model (Figure 1B), we used the GenomicSEM v0.0.2 package in R to conduct multivariate genome-wide association analyses of the latent genetic externalizing factor (EXT) and the residual genetic variance in ALCP (ALCP-S).

We first used the *sumstats* function to standardize the univariate GWAS summary statistics for each of the externalizing phenotypes. After excluding SNPs that were not present across all indicators, we retained a total of 6,132,068 SNPs for analysis at this stage. We then used the *userGWAS* function to perform the association test in an iterative manner for each SNP, where EXT and ALCP were both regressed onto the target SNP. These analyses were performed for SNPs that were available in all univariate summary statistics, had a minor allele frequency ≥ 0.5%, and were present in the 1000 Genomes Phase 3 (version 5) reference panel. As previously reported, we specified unit loading identification in these Genomic SEM models with SNP effects, where the factor loading of NSEX was fixed to 1 to set the scale of EXT. The field-standard threshold of $P < 5\times10^{-8}$ was used to designate genome-wide significance.

#### 2.2 Results

The results of the genome-wide association analyses for ALCP-O and ALCP-S are presented in **Supplementary Figures 1** and **2**, as Manhattan plots and quantile-quantile (Q-Q) plots, respectively. Briefly, we found robust polygenic signals for both ALCP-O and ALCP-S, as evidenced by substantial inflation of the test statistics ($\lambda_{\text{GC}} = 1.162$ and $\lambda_{\text{GC}} = 1.102$, respectively), LD Score regression intercepts (intercept = 1.013 and intercept = 0.990, respectively), and attenuation ratios (ratio = 0.068 and ratio = 0.000, respectively) for both phenotypes. Considered together, the observed inflation in test statistics appears to be due to a polygenic architecture rather than confounding or bias.

Before pruning for LD, we found that 542 SNPs were associated with ALCP-O at the level of genome-wide significance ($P < 5\text{x}10^{-8}$). As noted in the main text, 465/542 of these SNPs were genome-wide significant in EXT (~86%) and 60/542 were genome-wide significant in ALCP-S (~11%). There was no overlap between these two sets. Examination of the lead SNPs for ALCP-O in the EXT and ALCP-S results further emphasizes that our ‘residual GWAS’ framework was successful in delineating general versus specific genetic effects on problematic alcohol use (**Supplementary Table 1**).

#### 2.3 Genetic Correlations

We also used Genomic SEM to estimate genetic correlations between the latent genetic externalizing factor and complex traits that were broadly related to four domains: demography, health and medicine, psychopathology, and socioeconomic outcomes. We used a standard Benjamini-Hochberg false discovery rate correction (FDR 5%)[32] to account for multiple testing. Results for all phenotypes are presented in **Supplementary Table 2.**

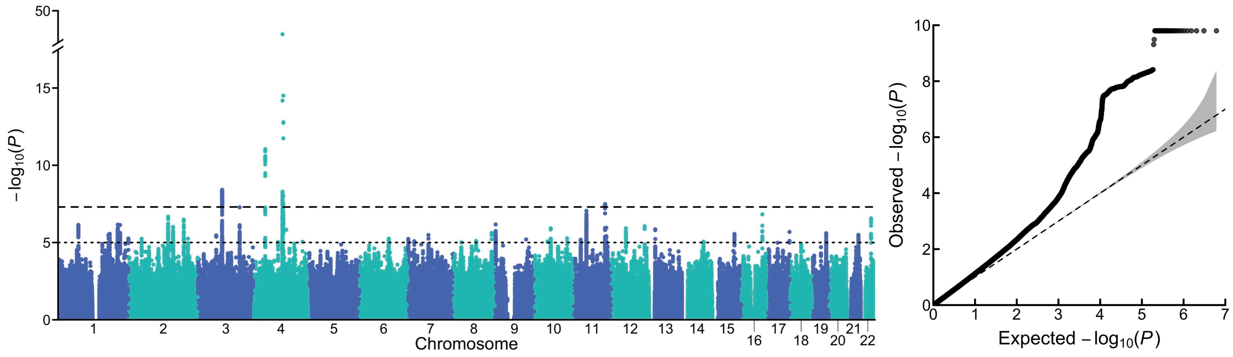


*Supplementary Figure 1: Manhattan Plot and QQ-plot for ALCP-O GWAS Results*

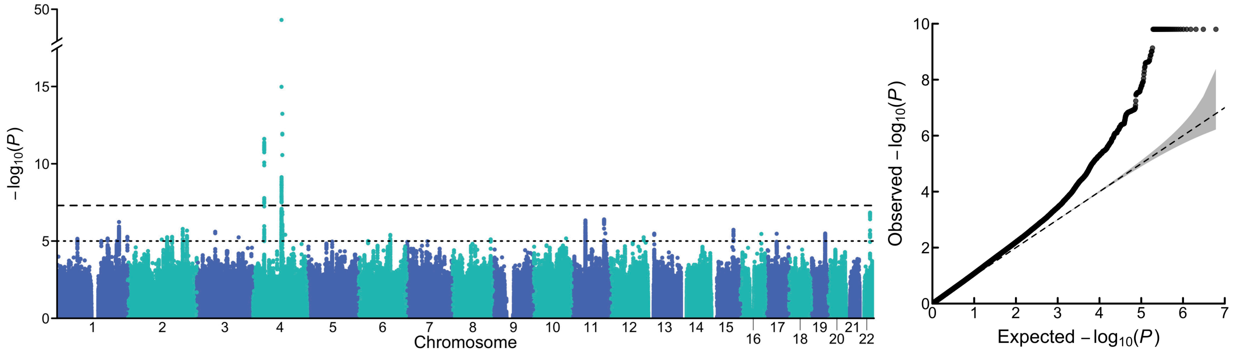


*Supplementary Figure 2: Manhattan Plot and QQ-plot for ALCP-S GWAS Results*

### 3 Bioannotation

In this section, we describe analyses comparing the biological function of results from the ALCP-O and ALCP-S GWASs, by applying several functional genomics tools to annotate and prioritize putative regulatory variants. These methods include functional annotations (i.e., CADD scores to identify highly deleterious SNPs), mapping annotations (i.e., eQTL SNP-gene expression association); as well as gene- and transcriptome-based analyses (MAGMA, H-MAGMA, and S-PrediXcan). Details of each method are presented below, and the results are reported in **Supplementary Tables 7-10**.

#### 3.1 Methods

##### *3.1.1 Functional mapping and annotation with FUMA*

We used Functional Mapping and Annotation of genetic associations (FUMA v1.3.5e)[33] to study the functional consequences of the lead SNPs from the ALCP-O and ALCP-S GWASs, reported in **Supplementary Table 1**, which included ANNOVAR categories (*i.e.*, the functional consequence of SNPs on genes), Combined Annotation Dependent Depletion (CADD) scores (*i.e.*, a measure of how deleterious a SNP is; greater than 12.37 is the suggested threshold to classify a SNP as deleterious), RegulomeDB scores (*i.e.*, a categorical score from 1a to 7 with 1a corresponding to the most biological evidence that the SNP is a regulatory element). The sources of the external reference data used in these analyses are fully described elsewhere[33].

##### *3.1.2 Gene-based, gene-set, and gene-property analyses with MAGMA*

We performed competitive gene-based association analyses using the genome-wide summary statistics from each GWAS by applying the method “multi-marker analysis of genomic annotation” (MAGMA v1.08)[33,34]. First, we assigned SNPs to genes based on physical position (gene-based analysis). SNPs were mapped to 18,318 protein-coding genes from Ensembl build 85. This approach uses multiple regression methods to account for LD between SNPs. All variants within all protein-coding genes were tested, using default settings, with LD structure estimated using the 1000 Genomes European sample as a reference. We evaluated Bonferroni-corrected significance, adjusted for testing 18,318 genes (one-sided $P < 2.73\times10^{-6}$). The results are reported in **Supplementary Table 7**.

Next, we performed a MAGMA gene-set analysis (the results are reported in **Supplementary Table 8**). We used 15,496 curated gene sets and Gene Ontology (GO) terms obtained from the Molecular Signatures Database (MsigDB v7.0, https://www.gsea-msigdb.org/gsea/msigdb/index.jsp)[35], which characterize the biological processes, molecular function and cellular component of individual gene products. We evaluated Bonferroni-corrected significance, adjusted for testing 15,496 gene sets (one-sided $P < 3.23\times10^{-6}$).

Lastly, we performed a gene property analysis to test the relationships between 54 tissue-specific gene expression profiles and gene associations (the results are reported in **Supplementary Figures 3-4**). We performed this analysis using the average expression of genes per tissue type as a gene covariate. Gene expression values were $\log_2$ transformed average RPKM (Reads Per Kilobase Million) per tissue type (after replacing RPKM > 50 with 50) based on GTEx RNA-seq data. We applied Bonferroni correction (one-sided $P < 9.26\times10^{-4}$) to correct for testing 54 gene expression profiles.

We used an extension of MAGMA v1.8, “Hi-C coupled MAGMA” or “H-MAGMA” [36], to assign non-coding (intergenic and intronic) SNPs to cognate genes based on their chromatin interactions. Exonic and promoter SNPs were assigned to genes based on physical position. We used four Hi-C datasets derived from adult brain[37], fetal brain[38], and iPSC derived neurons and astrocytes[39] (all available for download: https://github.com/thewonlab/H-MAGMA). The results are reported in **Supplementary Table 9**. We evaluated Bonferroni corrected *P*-value thresholds, adjusted for multiple testing within each analysis (one-sided $P < 9.84 \times 10^{-7}$, $P < 9.86 \times 10^{-7}$, $P < 9.84 \times 10^{-7}$, and $P < 9.83 \times 10^{-7}$, respectively).

###### *3.1.3 Gene-based association using transcriptomic data with S-PrediXcan*

We used S-PrediXcan v0.6.2[40] to analyze gene expression levels in multiple brain tissues, and to test whether the gene expression correlated with either ALCP-O or ALCP-S. The results are reported in **Supplementary Table 10**. We used pre-computed tissue weights from the Genotype-Tissue Expression (GTEx, v8) project database (https://www.gtexportal.org/) as the reference transcriptome dataset[41]. As input data, we used the summary statistics from each GWAS, transcriptome tissue data, and covariance matrices of the SNPs within each gene model (based on HapMap SNP set; available to download at the PredictDB Data Repository, http://predictdb.org) from 13 brain tissues: anterior cingulate cortex, amygdala, caudate basal ganglia, cerebellar hemisphere, cerebellum, cortex, frontal cortex, hippocampus, hypothalamus, nucleus accumbens basal ganglia, putamen basal ganglia, spinal cord and substantia nigra. We used a transcriptome-wide significance threshold of $P < 2.72 \times 10^{-7}$, which is the Bonferroni-corrected threshold when adjusting for 13 tissues times 14,117 tested genes (183,521 gene-tissue pairs).

#### 3.2 Results

###### *3.2.1 Results from the functional mapping and annotation with FUMA*

11 SNPs were genome-wide significantly associated with ALCP-O. The loci were located in 5 genomic regions spanning across 3 chromosomes, including the genes *CADM2*, *RNU6-887P*, *KLB*, *ADH1B*, *ADH1C*, *RP11-696N14.3*, *SLCC9A8*, *DRD2*. The SNPs spanning in these regions were primarily intronic (77.9%), 16.3% being intergenic and only 0.07% being exonic. On the other hand, 9 SNPs were genome-wide significantly associated with ALCP-S. All the loci were located on chromosome 4, and included the same genes identified in ALCP-O in this region (*KLB*, *ADH1B*, *ADH1C*, *RP11-696N14.3*, *SLCC9A8*). The majority of the SNPs identified with ALCP-S were intergenic (43%) or intronic (41%), a greater majority of the SNPs (2.7%) being exonic as compared to ALCP-O. Notably, the genes *CADM2* and *DRD2* were only implicated with ALCP-O but not ALCP-S.

###### *3.2.2 Results from the analyses with MAGMA, H-MAGMA, and S-PrediXcan*

In the MAGMA analysis, a total of 13 genes were found associated with ALCP-O at a Bonferroni-corrected significance (one-sided $P < 2.73 \times 10^{-6}$), including several novel genes that were not identified via GWAS, such as *MTCH2* (previously implicated with body mass index and other metabolic phenotypes[42,43]), *CELF1* (also previously implicated with body mass index[44]), *FNBP4* (previously associated with neuroticism[45] and nervousness[46]), among others. On the contrary, only *KLB* and *ADH1B* were associated with ALCP-S in this analysis. Full results are presented in **Supplementary Table 9**.

Tissue enrichment analyses for both ALCP-O and ALCP-S (**Supplementary Figures 3-4**) did not reach Bonferroni correction.

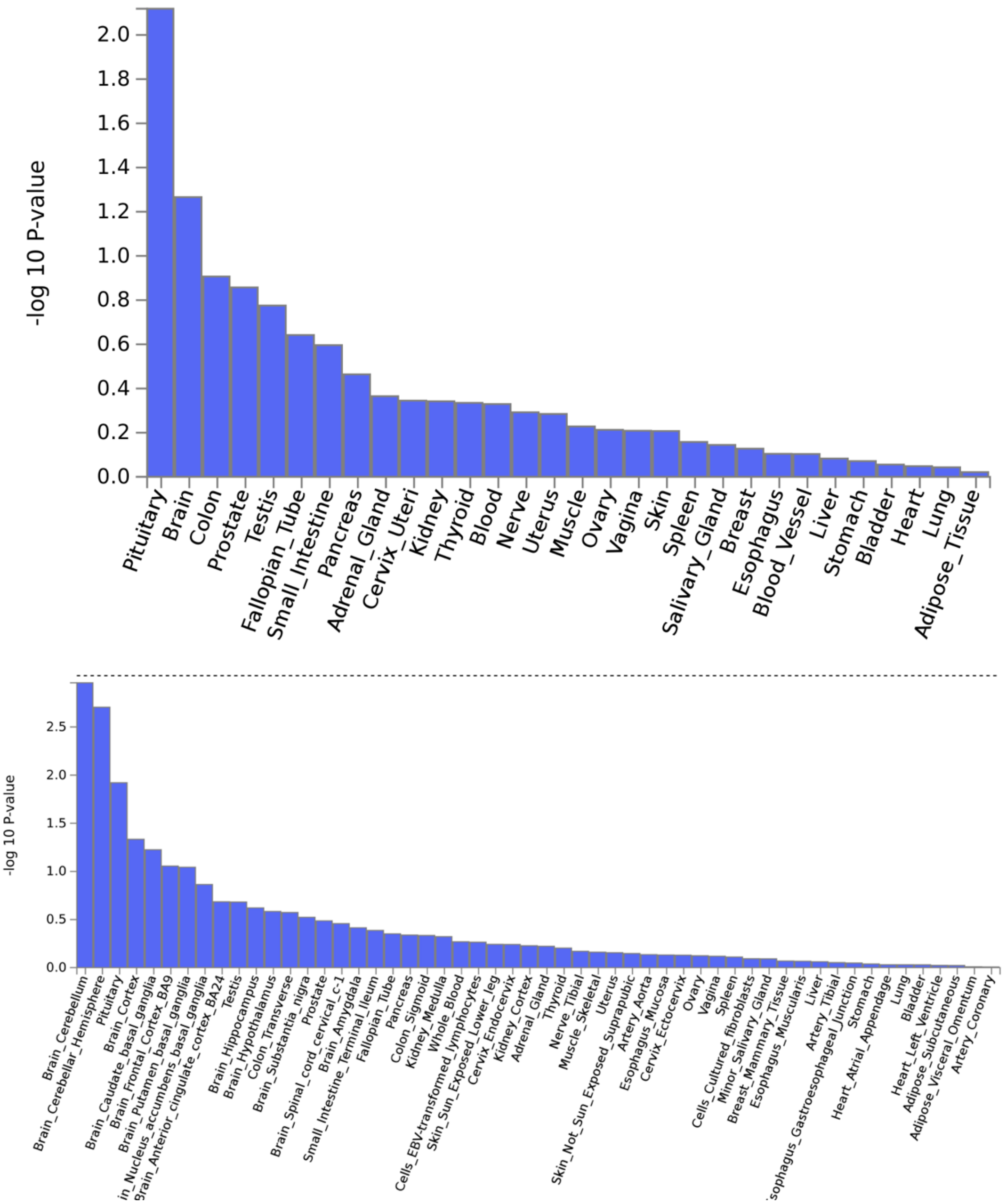


*Supplementary Figure 3: Gene property analyses results for ALCP-O*

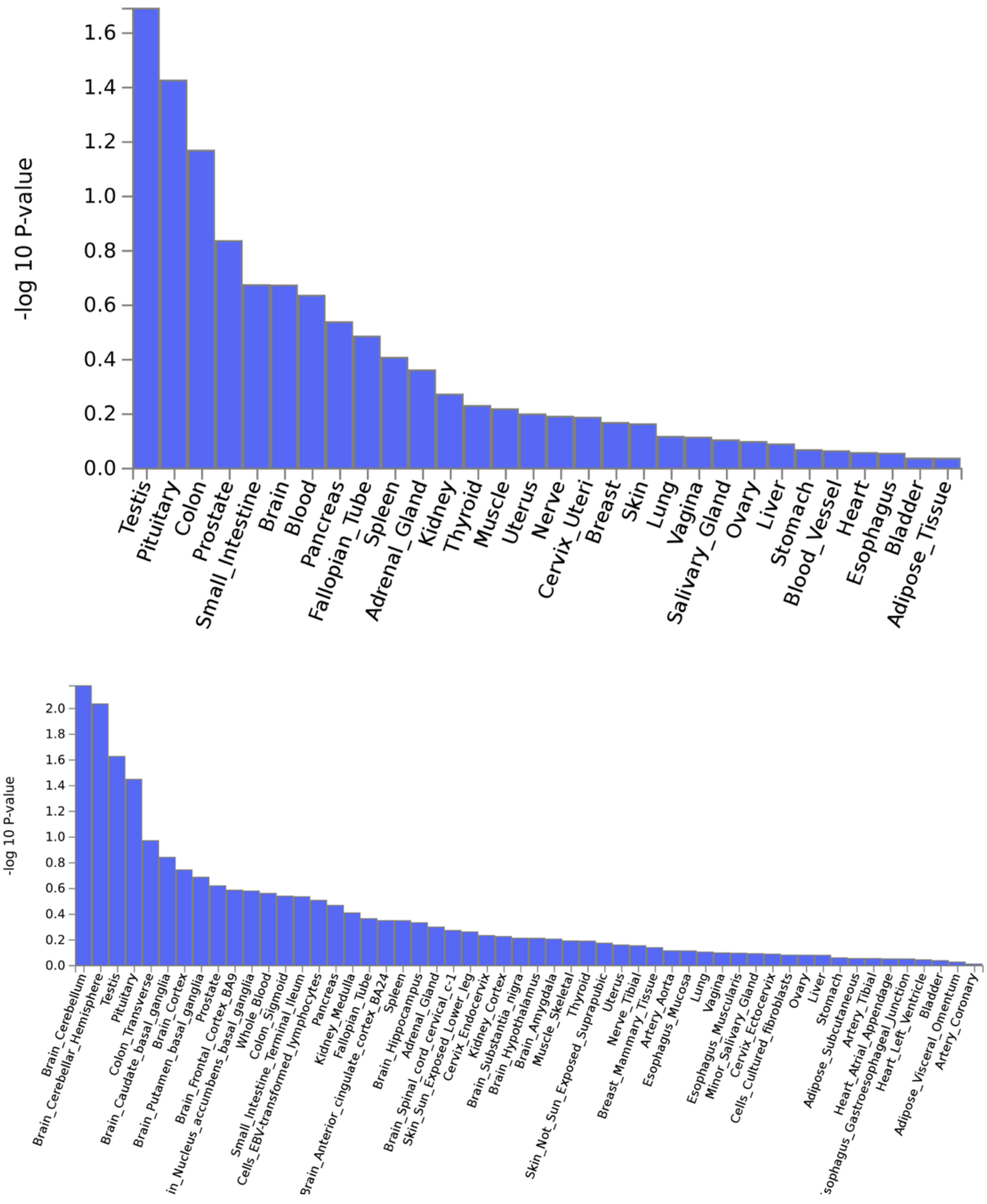


*Supplementary Figure 4: Gene property analyses results for ALCP-S*

Next, by analyzing gene regulatory relationships using H-MAGMA (**Supplementary Table 9**), we identified significant associations with ALCP-O in adult brain tissue (19 genes), fetal brain tissue (17 genes), iPSC-derived astrocytes (14 genes), and iPSC-derived neurons (17 genes). Of these 33 unique genes, the majority of the genes were also implicated in MAGMA gene-based analyses, including *DRD2*, *ADH1B*, *CELF1*, *FNBP4*, *MTCH2*, among others. In addition, H-MAGMA analyses implicated other ethanol metabolizing genes (*ADH4*, *ADH1A*), as well as other genes previously identified with alcohol, such as *TSPAN5*[14] or SLC39A13[47]. H-MAGMA also implicated other novel genes, such as *MIR5591*. Of these 33 genes, 29 were also implicated in the H-MAGMA analyses for ALCP-S, including *CADM2* or *DRD2*, which were not genome-wide significant in the GWAS, nor they were implicated in the MAGMA analysis. H-MAGMA analyses also revealed additional novel genes for ALCP-S, including WDR19 (previously nominally associated with cognitive function[48]).

Transcriptome-wide analyses with S-PrediXcan implicated *ADH1C* to be downregulated in the putamen (basal ganglia) both for ALCP-O and ALCP-S (**Supplementary Table 10**). S-PrediXcan analysis also identified and association between *NDUFS3* and ALCP-O, low cerebellar expression of *NDUFS3* being associated with ALCP-O (but not ALCP-S). *NDUFS3* has been previously associated with several other traits, including body size, insomnia, brain volume, loneliness, intelligence[49–55].

### 4 Polygenic Scores

In this section, we report on the construction and evaluation of polygenic scores (PGS) which we computed with weights from the problem alcohol use (ALCP), externalizing (EXT), and alcohol-specific (ALCP-S) GWASs. We compare the relative strength of each PGS using the incremental $R^2$ attained by adding the polygenic score to a regression model with baseline covariates, in accordance with previous efforts[14,24]. These analyses can be used to further refine pathways of risk for alcohol and other substance use disorders. Holdout Samples

#### 4.1 Samples

##### *4.1.1 The National Longitudinal Study of Adolescent to Adult Health (Add Health)*

Add Health is *nationally representative sample* with five waves of data collection spanning adolescence (ages 11-18 at baseline) through early midlife (ages 32-42 in the current follow-up). Add Health participants were selected from a stratified sample of 132 schools resulting in an initial, nationally representative sample of 90,118 students in grades 7-12. Of the original sample, 20,745 were selected for additional in-home interviews. Of those who completed the Wave I interview (1994-1995), 14,738 (71%) completed Wave II (1996); 15,197 (73%) completed Wave III (2001-2002); and 15,701 (75%) completed Wave IV (2007-2008). Most respondents completed the majority of the waves, with 16,278 (78%) completing three or more waves. Wave V (ages 32-42) data collection is underway, with a target sample of 19,828 (data for N=3872 is already released). In total, 15,159 individuals interviewed during Wave IV (ages 24-32) provided samples for genotyping, conducted using the Illumina Omni1 and Omni2.5 arrays. After quality control, genotypic data are available for 9,974 individuals (5,896 non-Hispanic White; 2,081 African American; 1,448 Hispanic; 550 Other).

##### *4.1.2 The Collaborative Study on the Genetics of Alcoholism (COGA)*

COGA, initiated in 1989 to identify vulnerability genes for AUD, ascertained *high-risk families* through adult probands in treatment for AUD. In the first 10 years, probands along with all willing first-degree relatives were assessed; recruitment was extended to include additional relatives in families that contained 2 or more first degree relatives with AUDs (n = 16,848). Data collection included a psychiatric interview, neurophysiological and neuropsychological protocols, and collection of blood for DNA. We currently have genome wide data on 12,145 individuals (8,038 individuals of European ancestry; 3,655 individuals of African ancestry). In 2004, COGA began the prospective study of adolescents and young adults, targeting assessment of youth aged 12-22 from COGA families where at least one parent had been interviewed. These subjects were re-assessed every two years; currently, 89% of individuals have 2+ interviews. COGA is *racially/ethnically diverse* (60.6% non-Hispanic White, 24.9% African American, 11.1% Hispanic, and 3.4% Other).

#### 4.2 Polygenic Score Creation

##### *4.2.1 Adjustment of GWAS effect sizes for linkage disequilibrium (LD)*

We adjusted GWAS effect sizes for the non-independence of nearby SNPs in the genome (referred to as linkage disequilibrium, or LD) using PRS-CS[56]. PRS-CS employs a Bayesian continuous shrinkage parameter to correct for LD using the 1000 Genomes European (1KG EUR) sample as a reference panel. In keeping with the recommendations of the developers, we utilized the 1KG EUR files distributed with the software and restricted to the ~1.3 million SNPs in the high-quality consensus genotype set defined by the HapMap 3 Consortium[56,57]. We generated polygenic scores using HapMap 3 SNPs that overlapped with the 1KG EUR panel and target samples (~1 SNPs in both COGA and Add Health). We applied the default parameters which allows PRS-CS to estimate the optimum global shrinkage parameter.

##### *4.2.2 Polygenic scores*

We computed polygenic scores (in individuals of European ancestries only[1]) from the weighted sum of the effect-coded alleles for a given individual *i*:

$$S_i = \sum_{j=1}^{M} \hat{\beta}_j g_{ij}$$

where $S_i$ is the polygenic score, $\hat{\beta}_j$ is the estimated additive effect of the effect-coded allele at SNP $j$, and $g_{ij}$ is the genotype at SNP $j$. The polygenic scores were standardized within each study cohort.

##### *4.2.3 Substance Use/Substance Use Disorder (SUD) Analyses*

Our choice of statistical model for the substance use/SUD analyses depended on (1) the distribution of the phenotype and (2) the structure of the data in the study cohort (independent vs. clustered or genetically related observations). In Add Health, the vast majority of the study participants are unrelated. Therefore, we analyzed one randomly drawn individual from any related pair (pairwise KING coefficient ≥ 0.0442). We used ordinary least squares (OLS) for continuous or ordinal outcomes, and logistic regression for binary outcomes. With OLS, we evaluated the traditionally defined coefficient of determination ($R^2$). In the case of logistic regression, we evaluated Nagelkerke's pseudo-$R^2$ [58]. COGA is a family-based cohort study with a variety of different pedigree structures[59–61]. To adjust for familial clustering, we utilized linear mixed models for continuous and ordinal outcomes (LMM) or generalized linear mixed models (GLMM) with a logistic link function for binary outcomes. For the LMM/GLMM models, we evaluated a pseudo-$R^2$ designed specifically for mixed models[62].

In Add Health (*N* = 5,107), for each phenotype ($Y$), we performed two regressions to estimate the accuracy of the polygenic score in explaining phenotypic variation. Specifically, we analyzed regression equations of the following form:

$$\textit{Baseline model:} \quad Y = X\beta + \varepsilon$$

$$\textit{Polygenic score model:} \quad Y = S\gamma + X\beta + \varepsilon$$

where $S$ and $X$ are matrices for the polygenic score and covariates with corresponding vectors of regression coefficients to be estimated, $\gamma$ and $\beta$, respectively. The baseline model

[1]Ancestry assignment was estimated from genetic data. See Braudt and Harris (2018) for full description of Add Health ancestry assignment. In COGA, ancestry was empirically assigned using the 1000 Genomes (phase 3) reference panel (YRI, CEU, JPT and CHB populations) as reference points[72].

included covariates for sex, age, and genetic principal components (PCs), as well as genotyping batch when applicable.

In COGA (*N* = 7,483), we estimated the following regression equations:

$$\textit{Baseline model:}\quad Y = X\beta + Z\mu + \varepsilon$$

$$\textit{Polygenic score model:}\quad Y = S\gamma + X\beta + Z\mu + \varepsilon$$

where we also included a matrix $Z$ with a binary indicator for each family unit and vector of unobserved random effects $\mu$ (specified as a variance component of the error term)[63]. The accuracy of the PGS was defined as the difference in $R^2$ (or pseudo-$R^2$) between the two models, a measure that is sometimes called "incremental $R^2$" (or $\Delta R^2$)[64].

##### *4.2.4 Longitudinal Growth Models*

To assess the associations between polygenic scores and change in alcohol-related behaviors over time, we fit a series of linear mixed-effects models with the data from four waves of Add Health, structured on age. In the notation of the multilevel model would appear as:

$$\textit{Level 1:}\quad Y_{ti} = \pi_{0i} + \pi_{1i} * \text{Age}_{ti} + \varepsilon_{ti}$$

$$\textit{Level 2 (intercept):}\quad \pi_{0i} = \beta_{00} + \Sigma_{k=1}\, \beta_{0k} * X_{ki} + \mu_{0i}$$

$$\textit{Level 2 (slope):}\quad \pi_{1i} = \beta_{10} + \Sigma_{k=1}\, \beta_{1k} * X_{ki} + \mu_{1i}$$

The model for within-individual change (Level 1) in AUI for time *t* of respondent *i* is composed of the respondents' baseline alcohol misuse ($\pi_{0i}$), the change in AUI as a function of age ($\pi_{1i}$), and any residual variance ($e_{ti}$). The Level 2 model, consisting of the between-individual portion (with a single, linear slope) includes the influence of *k* number of covariates (X's) on baseline values and change over time. Finally, we allow for random effects in the intercept and slope parameters ($u_{0i}$ and $u_{1i}$) which have the variance-covariance matrix, $\mu = \begin{bmatrix} \tau_1^2 & \tau_{21} \\ \tau_{12} & \tau_2^2 \end{bmatrix}$, and are assumed to ~$N(0,\ \mu)$ and uncorrelated with the fixed-effects in the model. The linear mixed model is well suited to handle longitudinal data that is unbalanced and/or has unique response schedule[65,66]. The model fitting process proceeded as follows:

1. Determine the functional form that the alcohol use index (AUI) followed over of time through both visual inspection and fitting linear and higher order polynomial models.
2. Determine the appropriate structure of the random effects and residual components of the model using nested models.
3. Examine whether polygenic scores for EXT and ALCP-S were associated with trajectories of AUI using both individual parameter estimates and improvements in overall model fit.

We included the effects of PGS and sex on both initial status and change over time because prior evidence of differences in trajectories across sex[67,68]. We also examined the possibility of sex differences by fitting sex-stratified models. All other covariates were included for their effect on initial status only to limit the number of parameters. The age categories at the ends of the age spectrum were collapsed due to small cell sizes (age ranges from 12 and below, to 32 and above).

#### 4.3 Phenotype definitions

##### *4.3.1 Substance use*

For substance use, we created measures of ever use for a variety of substances using each wave/interview in each sample. Respondents were classified as yes on *lifetime smoking initiation* if they responded yes to "Have you ever smoked cigarettes regularly, that is, at least 1 cigarette every day for 30 days?" at any point in Add Health, or yes to "Over your lifetime, have you smoked a total of 100 cigarettes (smoked 5 or more packs)?" at any point in COGA. The definition of *lifetime smoking initiation* and *cigarettes per day* in UKB has been described elsewhere[14]. *Lifetime alcohol use* was coded as yes if participants responded yes to "Have you had a drink of beer, wine, or liquor--not just a sip or taste of someone else's drink--more than 2 or 3 times in your life?" at any wave in Add Health or yes to either "Have you ever had a drink of alcohol?" or "So, you have never had even one full drink of alcohol?" in COGA. For cannabis use, participants were coded as a yes on *lifetime cannabis use* in Add Health or COGA if they responded yes to questions regarding ever using marijuana or hashish across any of the waves/interviews. Participants were classified as a lifetime opioid user (*lifetime opioid use*, COGA only) if they indicated opioids (among a list of many possible substances) for the question "Have you ever used any of these drugs to feel good or high, or to feel more active or alert? Or did you use any prescription drugs when they were not prescribed, or more than prescribed?" Finally, *lifetime other substance use* indicates whether participants have indicated they had ever used a variety of other illicit drugs or prescription medications outside their intended use. In Add Health this included ever using sedatives, tranquilizers, stimulants, painkillers, steroids, cocaine, crystal meth, and/or some other illicit substance. In COGA, the list of other substances included cocaine, stimulants, and/or sedatives.

##### *4.3.2 Substance use disorders*

In addition to substance use, we created measures of substance use disorders and/or problematic use, as the genetic overlap between use and problems is only partial[9]. Both Add Health and COGA contained some form of clinical interview[69,70]. We constructed measures substance use disorders that correspond to the substances as described above. In Add Health, *alcohol use disorder (AUD) symptoms*, *cannabis use disorder (CUD) symptoms*, and *other substance use disorder (other SUD) symptoms* were measured from the combined criteria counts of DSM-IV dependence and abuse of each of their corresponding substances. The total range of possible criteria ranged from 0 to 11. In COGA, *alcohol use disorder symptoms*, *cannabis use disorder symptoms*, *opioid use disorder (OUD) symptoms*, and *other substance use disorder symptoms* were measured from criteria counts of DSM-5 substance use disorder symptoms. These responses again ranged from 0 to 11. The only differences between combining abuse and dependence from DSM-IV criteria and using the DSM-5 criteria (which mostly reflects the combination of abuse and dependence into a single disorder) are in a single item. DSM-IV abuse contains the criteria of "[i]n the past year, have you more than once gotten arrested, been held at a police station, or had other legal problems because of your drinking?" which was not included in DSM-5. Instead, DSM-5 added, "[i]n the past year, have you wanted to drink so badly you couldn't think of anything else." Finally, in both Add Health and COGA, we used the Fagerstrom test for nicotine dependence (FTND) to assess *nicotine dependence symptoms*. The FTND assesses six criteria and has values ranging from 0 to 10. Overall, these measures of substance use disorders provide good coverage of problematic substance use.

##### *4.3.3 Alcohol Use Index (AUI)*

For the longitudinal growth models in Add Health, we created a composite measure of alcohol use/misuse across Wave I through Wave IV. This composite index included drinking frequency, drinking quantity, frequency of binge drinking, frequency of drinking to intoxication, and alcohol-related problems (e.g., problem with social relationships or commitments). Below are the specific questions and their responses:

1. *Drinking quantity* was assessed from the question: “Think of all the times you had a drink during the past 12 months. How many drinks did you usually have each time?”
2. *Drinking frequency* was assessed from the question: “During the past 12 months, how often did you drink beer, wine, or liquor?” Responses ranged from 0 = “Never” to 6 = “Every day or almost every day”.
3. *Binge drinking* was assessed from the question: “Over the past 12 months, on how many days did you drink 5 or more drinks in a row?” Responses ranged from 0 = “Never” to 6 = “Every day or almost every day”.
4. *Intoxication frequency* was assessed from the question: “During the past 12 months, on how many days have you been drunk or very high on alcohol?” Responses ranged from 0 = “Never” to 6 = “Every day or almost every day”.
5. *Alcohol Problems* was a composite assessed from the following questions at each wave (note: because the response options changed from Waves I-III to Wave IV, we recoded the outcomes, so each problem is coded 0=”Never”, 1=”Once”, and 2=”2+ times”):
    a. Wave I:
        i. “In the past 12 months: You got into trouble with your parents because you had been drinking”.
        ii. “In the past 12 months: You’ve had problems at school or with schoolwork because you had been drinking”.
        iii. “In the past 12 months: You had problems with your friends because you had been drinking”.
        iv. “In the past 12 months: You had problems with someone you were dating because you had been drinking”.
    b. Wave II:
        i. “In the past 12 months: You got into trouble with your parents because you had been drinking”.
        ii. “In the past 12 months: You’ve had problems at school or with schoolwork because you had been drinking.”.
        iii. “In the past 12 months: You had problems with your friends because you had been drinking.”.
        iv. “In the past 12 months: You had problems with someone you were dating because you had been drinking.”
    c. Wave III:
        i. “In the past 12 months: You had problems at school or work because you had been drinking”.

ii. “In the past 12 months: You had problems with your friends because you had been drinking”.

iii. “In the past 12 months: You had problems with someone you were dating because you had been drinking”.

iv. “How many times did you get into a sexual situation that you later regretted because you had been drinking?” or “How many times did you get into a physical fight because you had been drinking?” or “How many times did you show up to work/school drunk?”

d. Wave IV:

i. “How often has your drinking interfered with your responsibilities at work or school?”

ii. “How often have you been under the influence of alcohol when you could have gotten yourself or others hurt, or put yourself or others at risk, including unprotected sex?”

iii. “How often have you had legal problems because of your drinking, like being arrested for disturbing the peace or driving under the influence of alcohol, or anything else?”

iv. How often have you had problems with your family, friends, or people at work or school because of your drinking?”

All responses were coded so higher levels indicated greater levels of that alcohol-related behavior). Each of these items was then scaled to a value of 0 to 10 and averaged at each wave to create a composite measure ranging from normative use to problematic use. This method has been used in prior longitudinal models of alcohol use/misuse[71].

#### 4.4 Results of SUD analyses in Add Health and COGA

##### *4.4.1 Ever Substance Use*

Results for the PRS models are presented in **Supplementary Tables 3 & 4**. PRSs for ALCP-O were associated with cannabis use ($OR_{Add\ Health}$ = 1.15, $P = 7.14\text{x}10^{-6}$; $OR_{COGA}$ = 1.20, $P = 8.56\text{x}10^{-9}$), nicotine use ($OR_{Add\ Health}$ = 1.09, $P = 1.89\text{x}10^{-3}$; $OR_{COGA}$ = 1.23, $P = 2.35\text{x}10^{-11}$), other illicit substance use ($OR_{Add\ Health}$ = 1.20, $P = 2.24\text{x}10^{-8}$; $OR_{COGA}$ = 1.27, $P = 7.71\text{x}10^{-16}$), and opioid use ($OR_{COGA}$ = 1.24, $P = 4.09\text{x}10^{-10}$) in each of the samples in which they were measured, even after correcting for a false discovery rate of 5%. The only substance that the U-ALPCS PRS were not associated with was *lifetime alcohol use*, which is likely reflection of the ubiquitous nature of this phenotype (95–96% of the Add Health and COGA participants report lifetime alcohol initiation). When we compare this to results for ALCP-S, the PRSs for ALCP-S were only associated with cannabis use ($OR_{COGA}$ = 1.08, $P = 1.19\text{x}10^{-2}$), other illicit substance use ($OR_{Add\ Health}$ = 1.12, $P = 6.61\text{x}10^{-4}$; $OR_{COGA}$ = 1.15, $P = 1.83\text{x}10^{-6}$), and opioid use ($OR_{COGA}$ = 1.12, $P = 1.07\text{x}10^{-3}$) after correcting for multiple testing. In each case where ALCP-S remained significant, the effect sizes were markedly reduced. Finally, the EXT PRS were associated with every substance use phenotype in each of the samples, even after correcting for multiple testing (ORs = 1.32 – 1.70).

###### *4.4.1 Substance Use Disorder Criterion Count*

When we consider substance use disorder (SUD) criterion counts, we identified positive associations between the O-ALCP PRS and alcohol use disorder criteria (βAdd Health = 0.17, P = 3.53x10-7; βCOGA = 0.49, P = 4.62x10-29), cannabis use disorder criteria (βAdd Health = 0.11, P = 8.88x10-4; βCOGA = 0.16, P = 3.47x10-4), nicotine dependence symptoms (βCOGA = 0.20, P = 2.78x10-5), and other substance use disorder criteria (βCOGA = 0.20, P = 8.48x10-3) after correcting for multiple testing. The PRSs for ALCP-S were associated with alcohol use disorder criteria (βAdd Health = 0.13, P = 1.53x10-4; βCOGA = 0.34, P = 5.97x10-15), cannabis use disorder criteria (βAdd Health = 0.07, P = 3.17x10-2), and nicotine dependence symptoms (βCOGA = 0.10, P = 3.62x10-2). Again, these effect sizes are attenuated between O-ALCP and ALCP-S. If we use the more stringent Bonferroni threshold (.05/60, p < 0.000833333), only the association with AUD criteria remains. The PRS for EXT were significantly associated with each of the SUD criterion counts, even after correcting for multiple testing (β's = 0.11 – 0.59).

Finally, when we compare the variance explained by ALCP-O to that from adding the externalizing and the alcohol specific PGS (EXT + ALCP-S) jointly to the baseline model, we see an improvement in predictive power in both Add Health ($\Delta R^2_{EXT+ALCP-S}$ = 0.98%; $\beta_{EXT}$ = 0.20, $SE_{EXT}$ = 0.03, $P_{EXT}$ = 3.83x10-9; $\beta_{ALCP\text{-}S}$ = 0.14, $SE_{ALCP\text{-}S}$ = 0.03, $P_{ALCP\text{-}S}$ = 5.63x10-5) and COGA ($\Delta R^2_{EXT+ALCP-S}$ = 3.13%; $\beta_{EXT}$ = 0.58, $SE_{EXT}$ = 0.05, $P_{EXT}$ = 9.66x10-39; $\beta_{ALCP\text{-}S}$ = 0.35, $SE_{ALCP\text{-}S}$ = 0.04, $P_{ALCP\text{-}S}$ = 1.14x10-15), which was comparable to adding the ALCP-O and EXT PGS (Add Health $\Delta R^2_{EXT+ALCP-O}$ = 0.93%; COGA $\Delta R^2_{EXT+ALCP-O}$ = 3.21%). Because the PGS for EXT and ALCP-S are uncorrelated by design, the overall predictive power of these two scores is approximately the sum of the two. The overall predictive power of ALCP-O and EXT PGS is less than their sum as these two scores are correlated (r ~ 0.24 in both Add Health and COGA).

#### 4.5 Results of Longitudinal Growth Models in Add Health

##### *4.5.1 Initial model building*

We followed the steps for model building outlined in section 5.2.4, above. Results for the initial models are presented in **Supplementary Table 5**. We first ran an unconditional model (intercept only) to estimate the intraclass correlation coefficient (ICC), which provides an estimate of the degree of variance explained by between (vs within) cluster variance. For our alcohol use index (AUI), we estimated an ICC = .263, suggesting there is substantial within-person variance in AUI over time. In order to determine the functional form of change over time, we first plotted the mean levels of AUI by age and sex in the full AUI and each of its component indicators (see below).

For each of the AUI indicators and the AUI composite score, a quadratic change over time seemed to best describe the data. Including both linear ($\Delta\chi^2$ = 431.62, $\Delta$df = 1, $P$ = 7.22x$10^{-96}$) and quadratic ($\Delta\chi^2$ = 397.32, $\Delta$df = 1, $P$ = 2.11x$10^{-88}$) terms for age in the fixed effects portion of the model resulted in significantly better fit. We carried this model forward to estimate the structure of the random effects. Next, we evaluated several configurations of the random effects structure including random intercepts and slopes for the linear change in age (with a correlation between these two) and a random slope for both the linear and quadratic slopes of age (age and age$^2$). We settled on the latter due to the better fit indices (AIC = 55158.03 vs 55356.60) and the fact that the estimates for baseline values in AUI were very close to zero. We selected this quadratic model with random effects for age and age$^2$ as well as the covariance between the two) to test whether the EXT and ALCP-S PRSs were associated with baseline levels or change over time in AUI.

##### *4.5.2 Longitudinal Models of Polygenic Risk and Alcohol Misuse*

Using the above model as our baseline model, we tested the association between polygenic scores for EXT and ALCP-S with baseline levels and change over time in AUI. For each model, we included sex and the first ten ancestral principal components (PCs) as covariates. Beginning with EXT, we tested the associations in a stepwise approach, first adding the EXT PRS in the first model, and then interaction between the EXT PRS and age, followed by the interaction between the EXT PRS and age$^2$. Including the EXT PRS in the model for baseline AUI significantly improved the fit of the model ($\Delta\chi^2$ = 69.20, $\Delta$df = 1, $P$ = 8.89x$10^{-17}$), while including the EXT PRS in the model for linear change ($\Delta\chi^2$ = 0.49, $\Delta$df = 1, $P$ = 4.82x$10^{-1}$) or quadratic change ($\Delta\chi^2$ = 0.02, $\Delta$df = 1, $P$ = 8.87x$10^{-1}$) did not improve the fit of the model. We therefore only carried forward the effect of EXT PRS on baseline AUI in the models testing the ALCP-S PRS and followed the

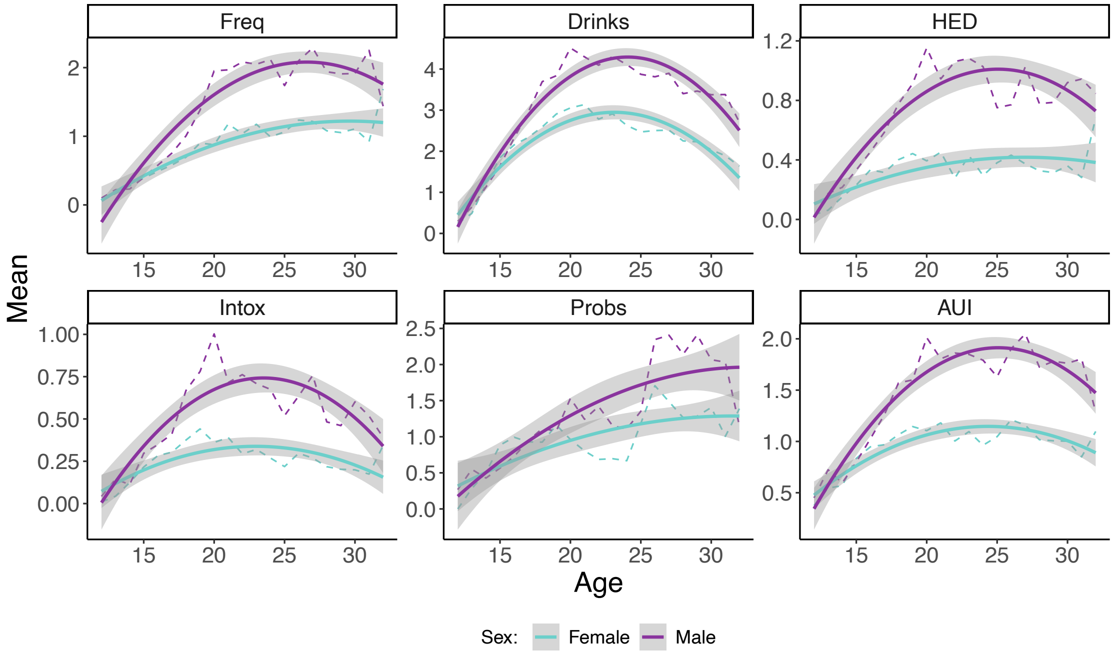


*Supplementary Figure 5 Mean Levels of Alcohol Phenotypes Over Time in Add Health*

Mean levels of drinking frequency (Freq), drinking quantity (Drinks), binge drinking (HED), drinking to intoxication (Intox), alcohol-related problems (Probs), and alcohol use index (AUI) from ages 12 to 32 in Add Health (N = 5,107).

same stepwise approach. ALCP-S significantly improved fit for both the baseline ($\Delta\chi^2$ = 13.48, $\Delta$df = 1, $P$ = 2.41x10$^{-4}$) and linear change ($\Delta\chi^2$ = 19.59, $\Delta$df = 1, $P$ = 9.59x10$^{-6}$) models. The overall model that best describes the relationship between the EXT and ALCP-S PRSs and change in AUI over time is one in which those with higher levels of EXT have greater levels of AUI at baseline, and those with higher ALCP-S have both a greater AUI at baseline and a steeper increase in AUI over time.

The final tests we performed were to assess potential sex differences in trajectories of AUI, especially given the differences in mean levels seen in plotting the observed values. We tested two potential models for sex differences. In the first model, we allowed the effect of change over time to vary by sex (e.g., inclusion of sex-by-age and sex-by-age$^2$ interactions). In the second model we also allowed the effect of each PRS to vary by sex (e.g., inclusion of sex-by-PRS, sex-by-PRS-by-age, and sex-by-PRS-by-age$^2$ interactions). Full parameter estimates for these models are available in **Supplementary Table 6**. Including sex differences in change over time resulted in a substantial increase in model fit ($\Delta\chi^2$ = 244.58, $\Delta$df = 2, $P$ = 7.77x10$^{-54}$). Including the interactions with PRS also significantly improved model fit ($\Delta\chi^2$ = 18.28, $\Delta$df = 3, $P$ = 3.85x10$^{-4}$). However, given that only one of the interactions was significant (sex*age*ALCP-S PRS), and the relatively marginal significant of this interaction ($\beta$ = 0.070, $P$ = 4.58x10$^{-2}$), we did not select the model with full sex differences and instead selected the first model with sex differences in change over time, only. The figure below provides predicted trajectories across sex, PRS (± 1.5 SD of both EXT and ALCP-S PRS), and age.

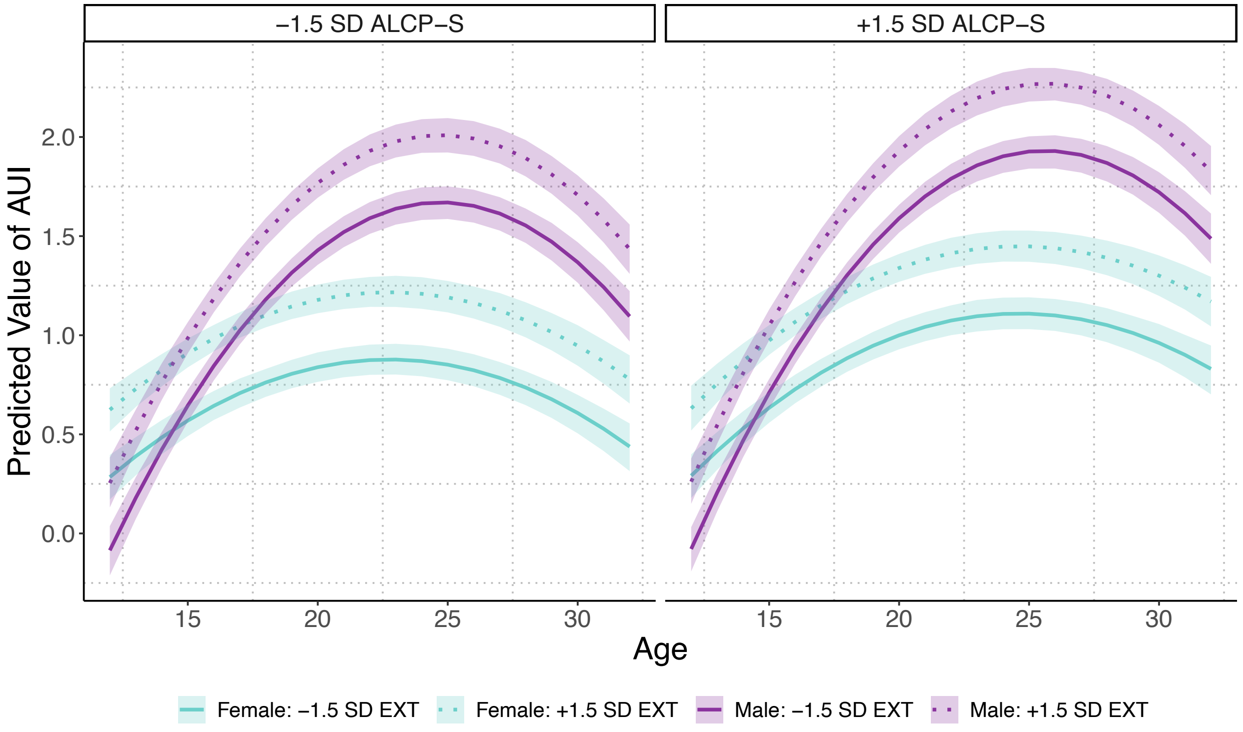


*Supplementary Figure 6: Predicted Trajectories of AUI Across Age, Sex, and Polygenic Risk*

Predicted values for alcohol use index (AUI) from ages 12 to 32 using linear mixed models in Add Health (N=5,107). Values for EXT and ALCP-S polygenic scores set to ±1.5 SD. The shaded areas represent 95% confidence intervals. Confidence intervals estimated using percentile method bootstrapping over 1000 bootstrap samples. All other covariates set to mean values.
